# Racial Disparities in the Utilization and Outcomes of concomitant Left Atrial Appendage Exclusion during Cardiac Surgery: A Nationwide Analysis

**DOI:** 10.1101/2024.03.05.24303836

**Authors:** Izhan Hamza, Mahmoud Ismayl, Amer Abdulla, Patricia A. Pellikka, Khaled Chatila

## Abstract

**Background:** Concomitant left atrial appendage exclusion (LAAE) during cardiac surgery is an effective method of reducing stroke risk in patients with atrial fibrillation. Previous studies have documented racial disparities in the performance of multiple cardiac procedures. We sought to evaluate if there were any disparities in the utilization and outcomes of concomitant LAAE.

**Methods:** We used the 2016-2020 National Inpatient Sample database to identify hospitalizations for cardiac surgery in patients with atrial fibrillation with concomitant LAAE. We utilized the weighted data to compare the in-hospital mortality and complications such as stroke, bleeding, infection, heart failure and pericardial complications among different race/ethnic groups.

**Results:** From 2016 to 2020, 432,244 hospitalizations were for cardiac surgery, of which 91,395 (21%) included concomitant LAAE. Of these, 77,440 (84.7%) were in White patients, 4,179 (4.6%) were in Black patients, 4,834 (5.3%) were in Hispanic patients, and 4,939 (5.4%) were in other races. Black and Hispanic patients had lower odds of undergoing concomitant LAAE during cardiac surgery compared to white patients (adjusted odds ratio (aOR) 0.85, 95% confidence interval (CI) 0.79-0.93 and aOR 0.85, 95% CI 0.79-0.93, respectively). There were no significant differences between Black and Hispanic patients in in-hospital mortality or procedural complications except for higher bleeding complications in Hispanic patients (aOR 2.38, 95% CI 1.27-2.86) compared to White patients. Through the study period, the proportion of patients receiving concomitant LAAE increased in all race/ethnic groups.

**Conclusion:** Concomitant LAAE during cardiac surgery is underutilized in Black and Hispanic patients compared to White patients, despite mostly similar clinical outcomes. Further comparative longitudinal studies are warranted to confirm these findings.

## Introduction

Stroke is a devastating complication in patients who suffer from atrial fibrillation (AF) (1). Most of the clots leading to cardioembolic stroke are thought to originate in the left atrial appendage (LAA) (2) In order to mitigate the risk of stroke in patients with AF, present guidelines recommend either the use of anticoagulation or closure of the LAA (2,3). The Closure of the LAA can be achieved both percutaneously and surgically (4,5). The Left Atrial Appendage Occlusion during Cardiac Surgery to Prevent Stroke (LAOOS III) trial was the first randomized controlled trial demonstrating the clear benefit of surgical closure of LAAE in patients with AF (6). As a result, the 2023 AF guidelines have changed the strength of recommendation for concomitant LAAE during cardiac surgery from Class 2b to Class 1a (7) (8,9). However, the performance of LAAE during cardiac surgery continues to depend on region and institution practices (10).

Previously, racial/ethnic disparities in the utilization and outcomes of multiple cardiac procedures and therapies have been documented and studied (11–14). However, racial/ethnic disparities in the utilization and outcomes of concomitant LAAE during cardiac surgery for AF patients have not been evaluated. Therefore, we sought to evaluate for the presence of any racial/ethnic disparities in the choice to exclude the LAA and in the outcomes of LAAE.

## Methods

### Data source

The National Inpatient Sample (NIS) was utilized to extract relevant hospitalizations from 2016 through 2020. The NIS is a nationwide administrative database developed by the Agency for Health Care Research and Quality (AHRQ)(15). It contains a stratified unweighted sample of 20% of all national hospitalizations using state-level data. The discharge data includes data from over 4000 hospitals from over 46 states, accounting for 96% of all hospital discharges. After weights are applied, the NIS corresponds to approximately 35 million hospital discharges. The NIS database contains patient information such as age, sex, race, primary and secondary diagnosis, procedures performed, insurance status, hospitalization outcome, length of stay, and total cost.

### Patient selection

We used *International Classification of Diseases, tenth revision (ICD-10)* diagnosis codes to identify hospitalizations in which adult patients with AF underwent cardiac surgery (defined as coronary artery bypass grafting [CABG] or any valvular surgery) with concomitant surgical LAAE and stratified them based on race/ethnicity. Similar to prior NIS studies, we used four race/ethnic groups (White, Black, Hispanic, and Other), races/ethnicities with a smaller sample size (Asian, Pacific Islander, American Indian or Alaska Native, and Other) were combined as other. (16) . NIS combines “race” and “ethnicity” into 1 data element (race). If both “race” and “ethnicity” were available, ethnicity was preferred over race in assigning the HCUP value for “race” (17). All ICD-10 codes used in this study can be found in the *Supplemental material Table 1*. We excluded patients under 18 years of age, those with missing information on race/ethnicity, and those who had a prior CABG or valve surgery. A study flow diagram is shown in *Figure 1*.

**Figure 1.**
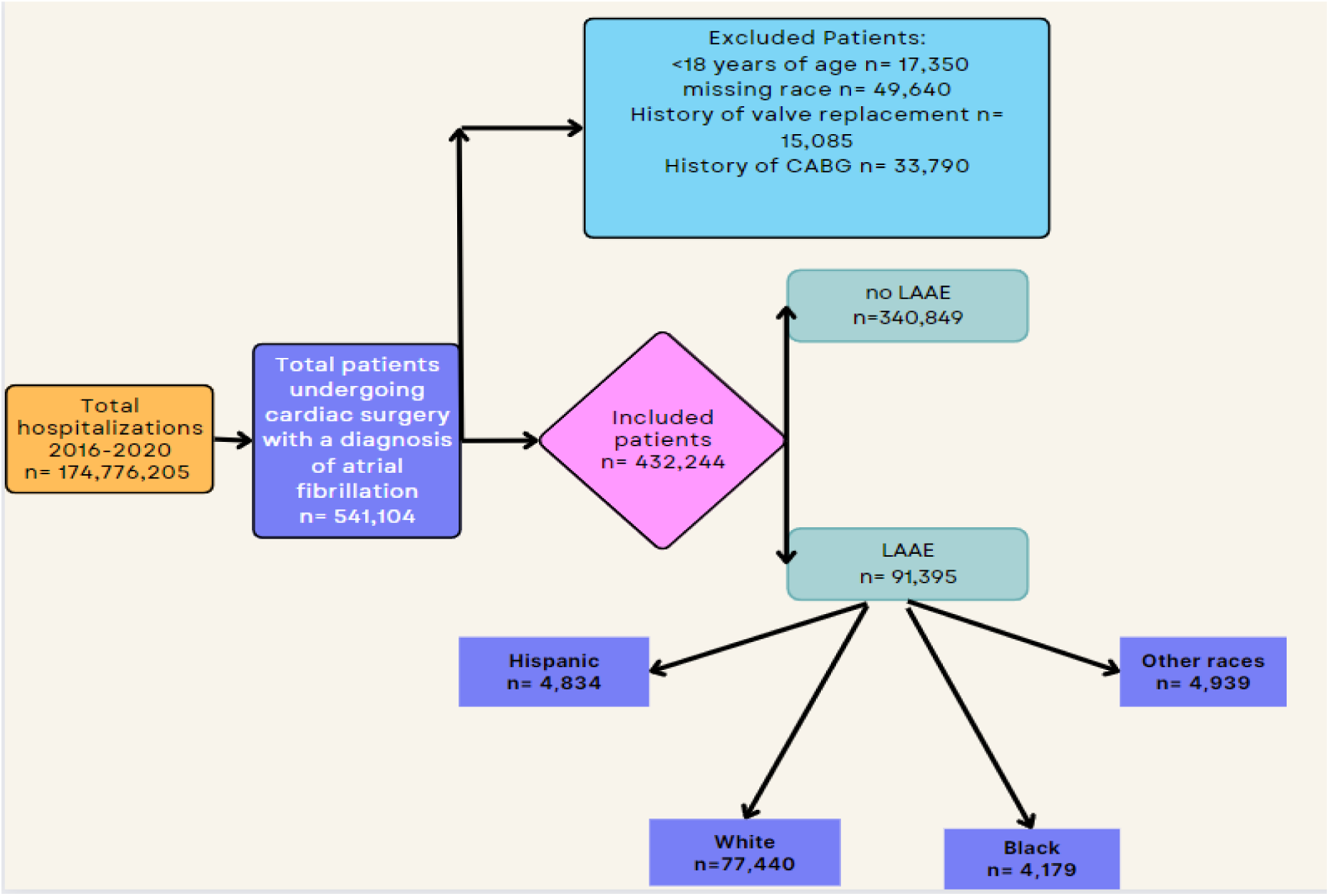
Study flow showing study inclusion and exclusion criteria.

**Table 1.**
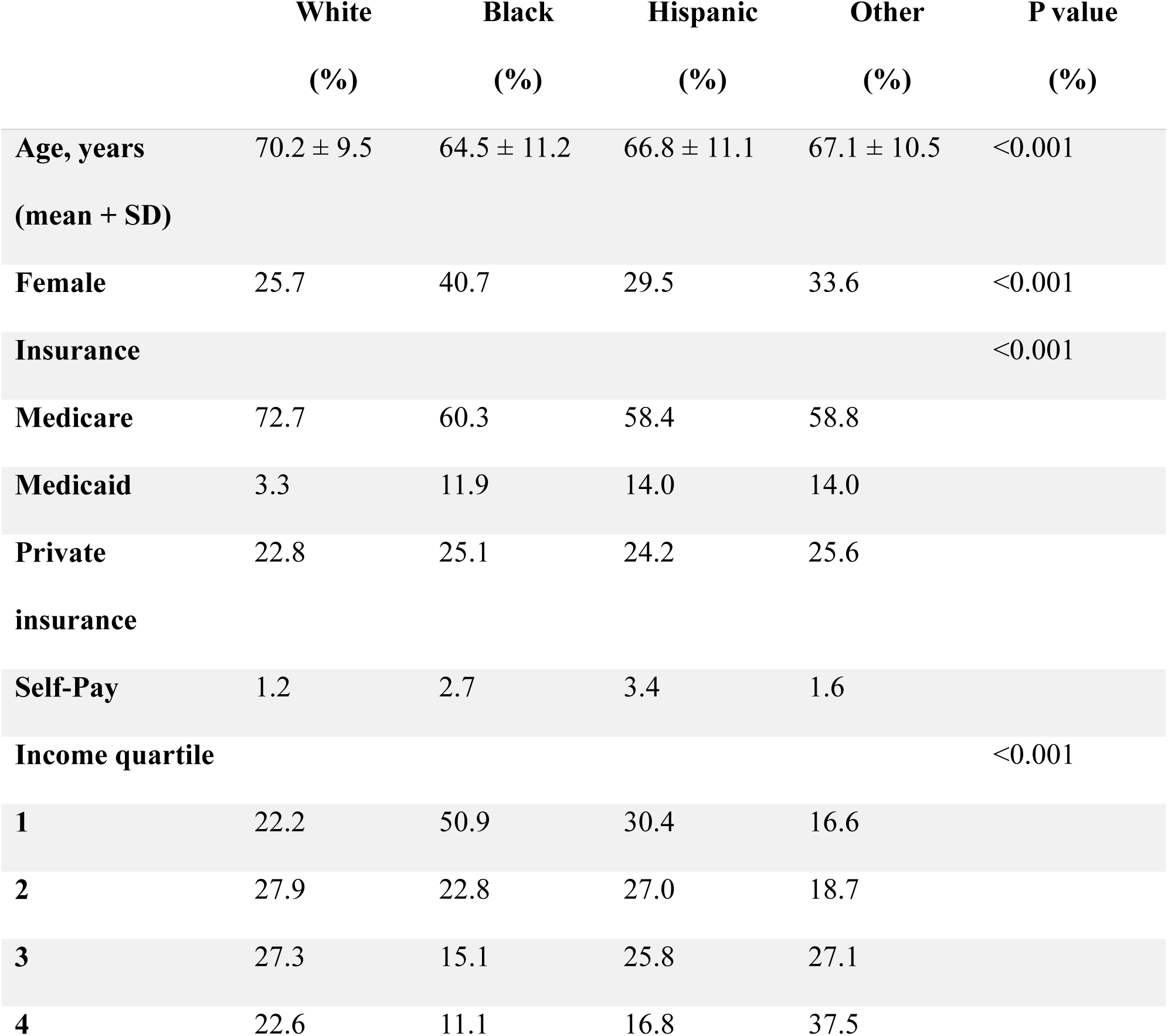
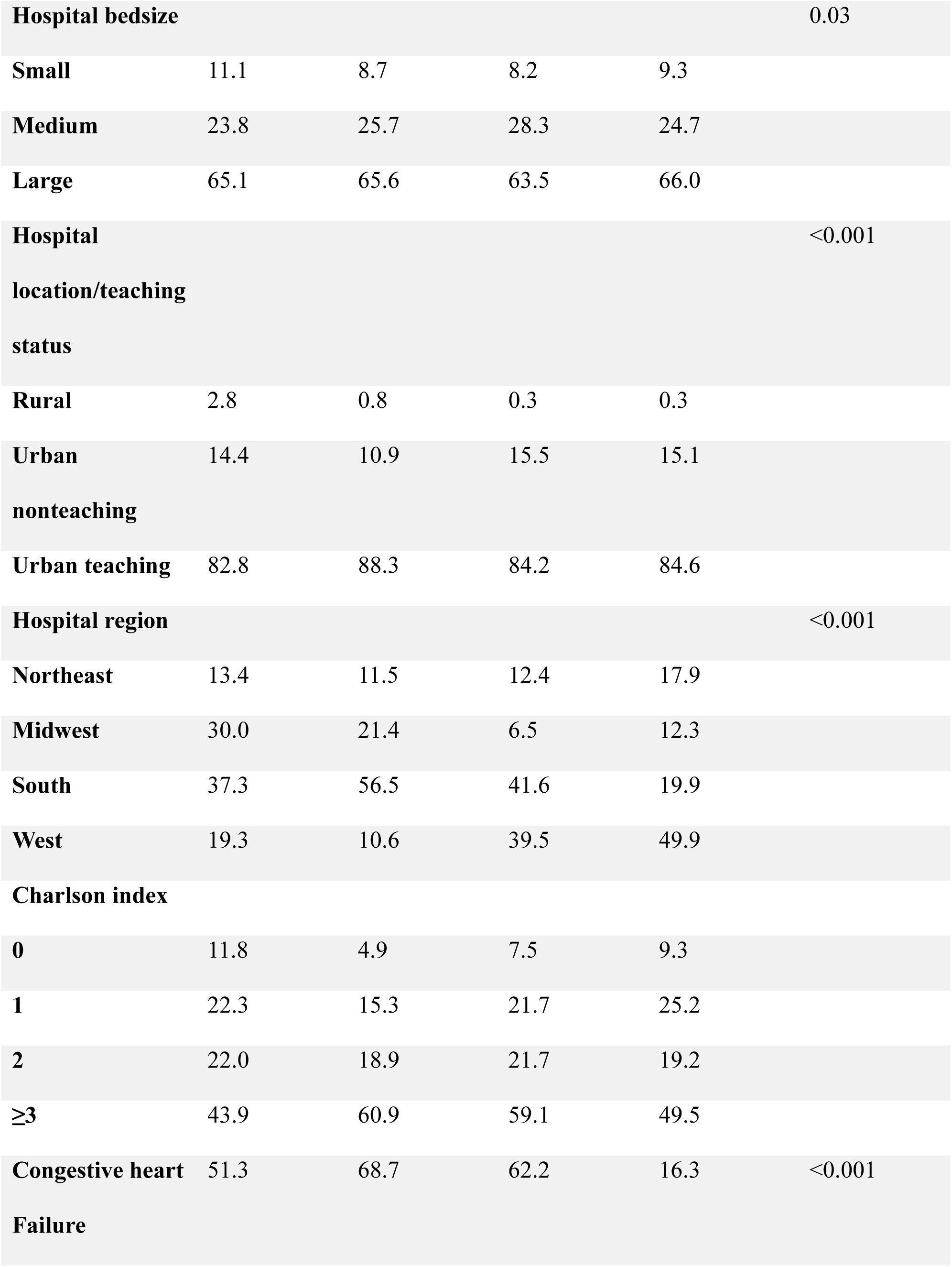
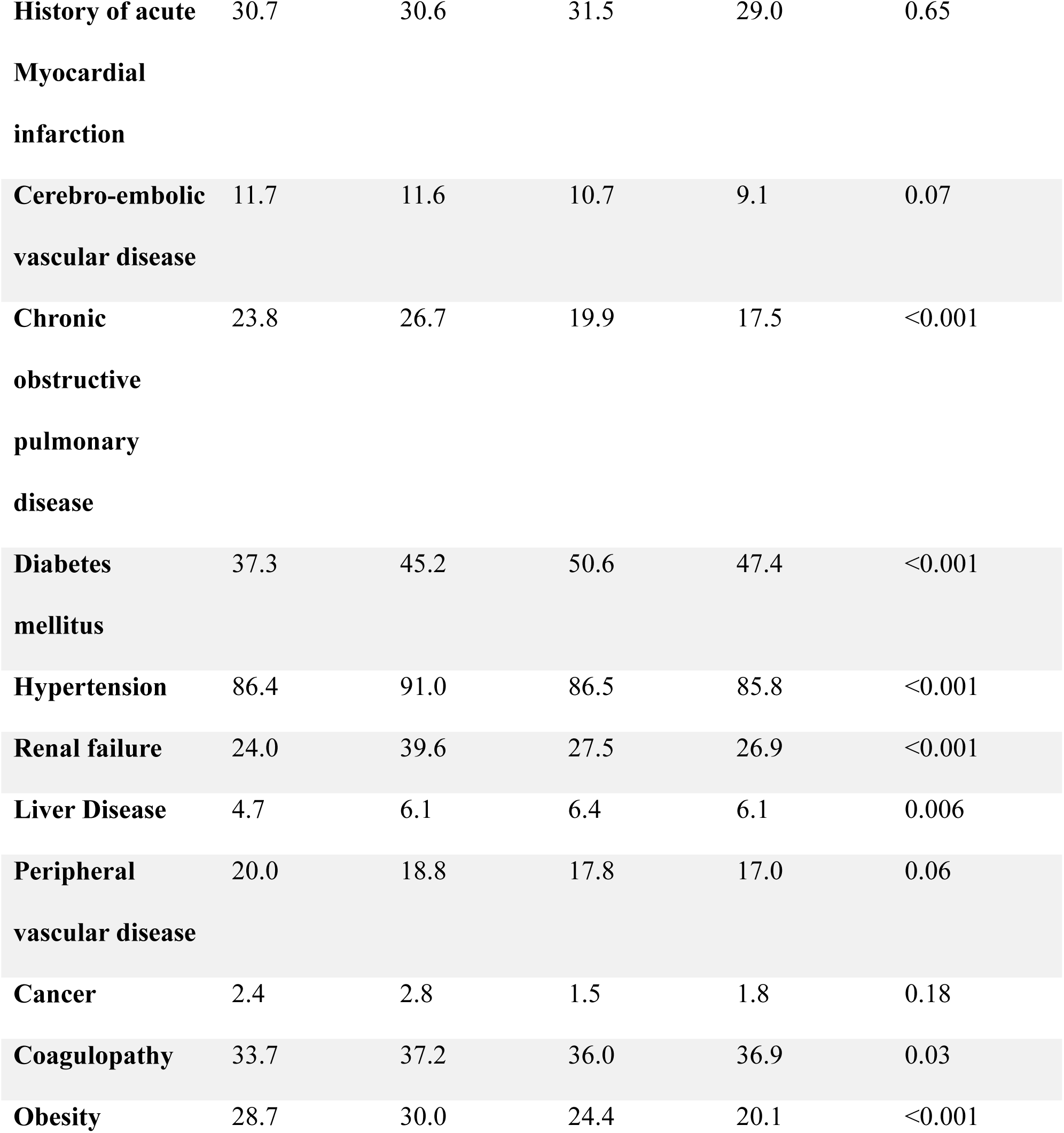
Patient demographic, clinical and hospital level characteristics based on race. More information on hospital bed size and income quartile parameters can be found on (https://www.hcup-us.ahrq.gov/db/nation/nis/nisdde.jsp). Other races refers to Asian, Pacific Islander, American Indian or Alaska Native, and other.

### Outcomes

The primary outcome was in-hospital all-cause mortality, while secondary in-hospital outcomes included stroke, bleeding complications, pericardial complications, procedure-related infections, hospital length of stay (LOS), and total hospital charges.

### Statistical analysis

Categorical variables were compared using Chi-square test, while continuous variables were compared using linear regression or paired t-test. We reported categorical variables as percentages and continuous variables as means with their associated standard deviations (SD).

For outcome analysis, a multivariable logistic and linear regression model was applied where appropriate, utilizing baseline and hospital-level characteristics to evaluate the impact of concomitant LAAE on primary and secondary outcomes as well as the odds of receiving LAAE during cardiac surgery based on race/ethnicity. Adjustment variables included in the multivariable regression analysis were selected after performing a univariable screen and after a careful literature review. Variables that would not have been included per univariable screen but were known confounders were forced into the multivariable analysis. Variables adjusted for included hospital region and size, patient income quartile, insurance type, Charlson comorbidity scores, age, gender, and other relevant comorbidities (Supplemental material Table 2-9).

**Table 2.**
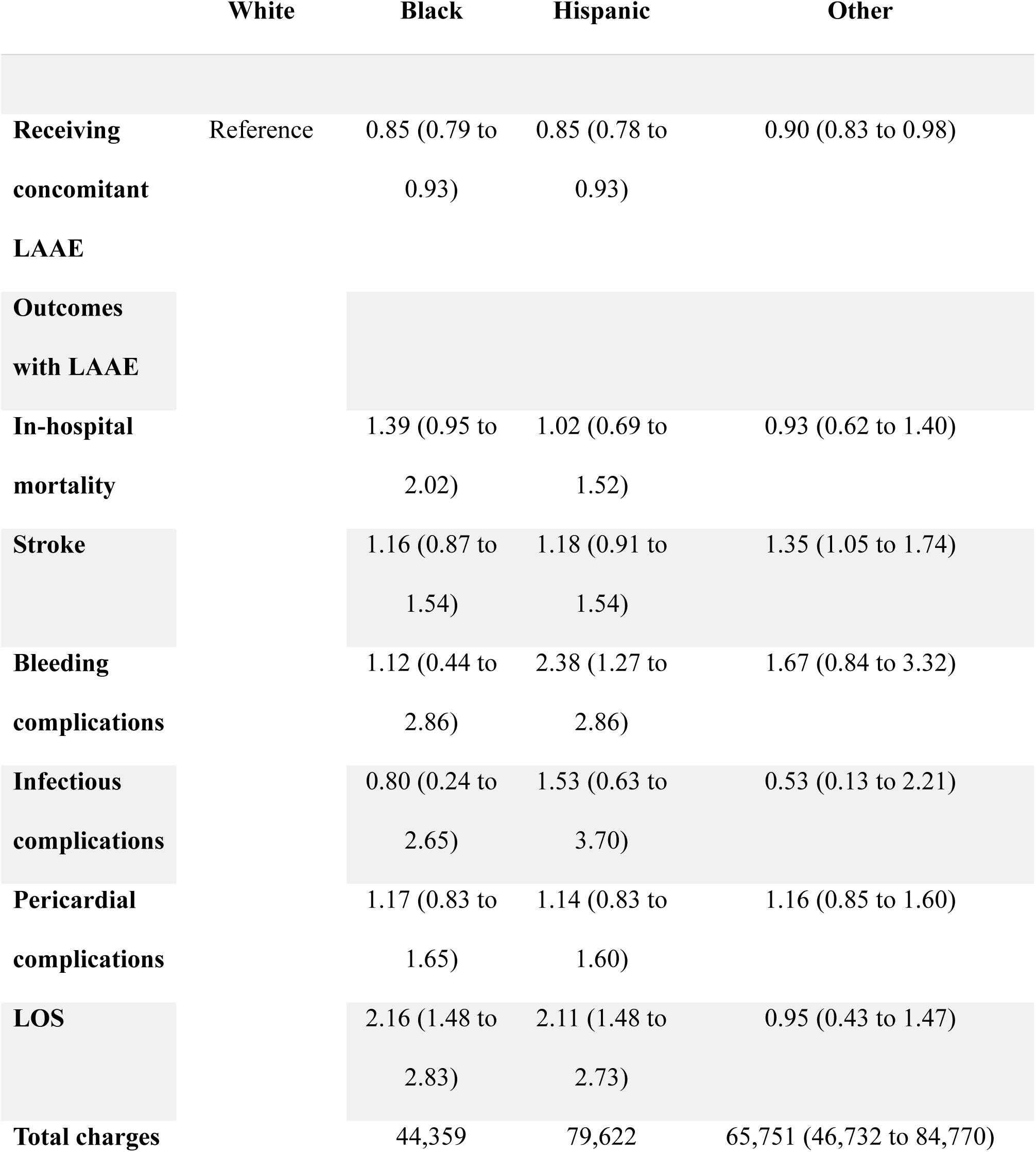
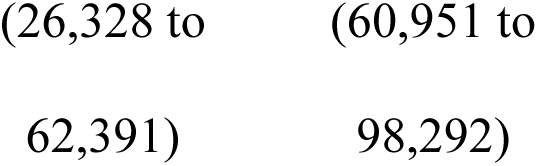
LAAE Utilization rates and patient clinical outcomes based on patient race/ethnicity; Values are expressed as adjusted odds ratios with their respective 95 % confidence intervals.

All analyses were performed in compliance with the HCUP user agreement and regulations. Analyses were performed using Stata version 18 (StataCorp, College Station, Texas) software utilizing the NIS sample weights provided by HCUP to obtain national estimates. We used “<11” for variables with a small number of observations, which would pose a risk to patient privacy violations. A two-tailed p-value of < 0.05 was considered statistically significant.

## Results

### Baseline characteristics

From 2016 through 2020, a total of 91,395 hospitalizations for concomitant LAAE during cardiac surgery were included in our study. Of these, 77,440 (84.7%) were in White patients, 4,179 (4.6%) were in Black patients, 4,834 (5.3%) were Hispanic, and 4,939 (5.4%) were in other race/ethnic groups.

Black patients were more likely to be female, younger, uninsured, and belong to lower income quartiles compared to the other races/ethnicities. Additionally, Black patients were more likely to have congestive heart failure, chronic obstructive pulmonary disease, hypertension, renal disease, coagulopathy, and obesity compared to other races/ethnicities. On the other hand, Hispanic patients were more likely to be diabetic and have liver disease compared to the other races/ethnicities. A full breakdown of patient, hospital, and clinical characteristics stratified by race/ethnicity is shown in *Table 1*.

### Racial disparities in receiving LAAE and clinical outcomes

Among patients undergoing cardiac surgery, Black race and Hispanic ethnicity were independently associated with lower odds of receiving concomitant LAAE (adjusted odds ratio [aOR] 0.85, 95% confidence interval [CI] 0.79-0.93 for Black race and aOR 0.85, 95% CI 0.79-0.93 for Hispanic ethnicity, *see Table 2*).

In-hospital mortality was similar in non-White races and White races. The odds for in-hospital mortality was 1.39 (95% CI 0.95 to 2.02) for Black patients, 1.02 (95% CI, 0.69 to 1.52) for Hispanics, and 0.93 (95% CI 0.62 to 1.40) for other races compared to White patients. Both Black and Hispanic patients had similar odds of stroke compared to White patients (aOR 1.16, 95% CI 0.87-1.54 for Black patients and aOR 1.18, 95% CI 0.91-1.54 for Hispanic patients). However, other races had higher odds of stroke compared to white patients (aOR 1.35, 95% CI 1.05-1.74). Black and other races have similar odds of bleeding complications compared to white patients (aOR 1.12, 95% CI 0.44-2.86 for Black patients and aOR 1.67, 95% CI 0.84-3.32 for other races). However, Hispanic patients had higher odds of bleeding complications (aOR 2.38 95%, CI 1.27-2.86) compared to White patients. Infectious complications were similar between white and non-White races (aOR 0.80 95%CI 0.24-2.65 for Black patients, aOR 1.53, 95% CI 0.63-3.70 for Hispanic patients, and aOR 0.53, 95% CI 0.13-2.21 for other races compared with White race). Lastly, pericardial complications were similar between White and non-white races (aOR 0.80, 95% CI 0.24-2.65 for Black patients, aOR 1.53, 95% CI 0.63-3.70 for Hispanic patients, and aOR 1.16, 95% CI 0.85-1.60 for other races compared with White patients).

### LOS and Total Hospital Charges

For patients undergoing concomitant LAAE, non-white races had longer LOS compared to white patients. The mean difference in LOS was 2.16 days (95% CI 1.48-2.83) for Black patients, 2.11 days (95% CI 1.48-2.73) for Hispanic patients, and 0.95 days (95%CI 0.43-1.47) for other races/ethnicities compared to White patients. Non-white races also had higher total hospital charges compared to White patients. The mean difference in total hospital charges was $44,359 (95% CI $26,328-$62,391) for Black patients, $79,622 (95% CI $60,951-$98,292) for Hispanic patients, and $65,751 (95% CI $46,732-$84,770) for other races compared to White patients.

### Yearly Trend in LAAE by race

Over the 5-year study period, the trend in LAAE has increased in all race/ethnic groups. The proportion of White patients undergoing concomitant LAAE increased from 14.8% to 19.8% (Ptrend <0.001), and the proportion of Black and Hispanic patients undergoing concomitant LAAE increased from 14.4% to 21.2% and 15.3% to 22.1% (Ptrend <0.001 for both), respectively, See *Figure 2*. However, there was no difference in the yearly increase in concomitant LAAE when comparing White and non-White races, See *Table 3*.

**Figure 2.** Year on year trend of proportion of patients undergoing LAAE based on race/ethnicity.

**Table 3.**
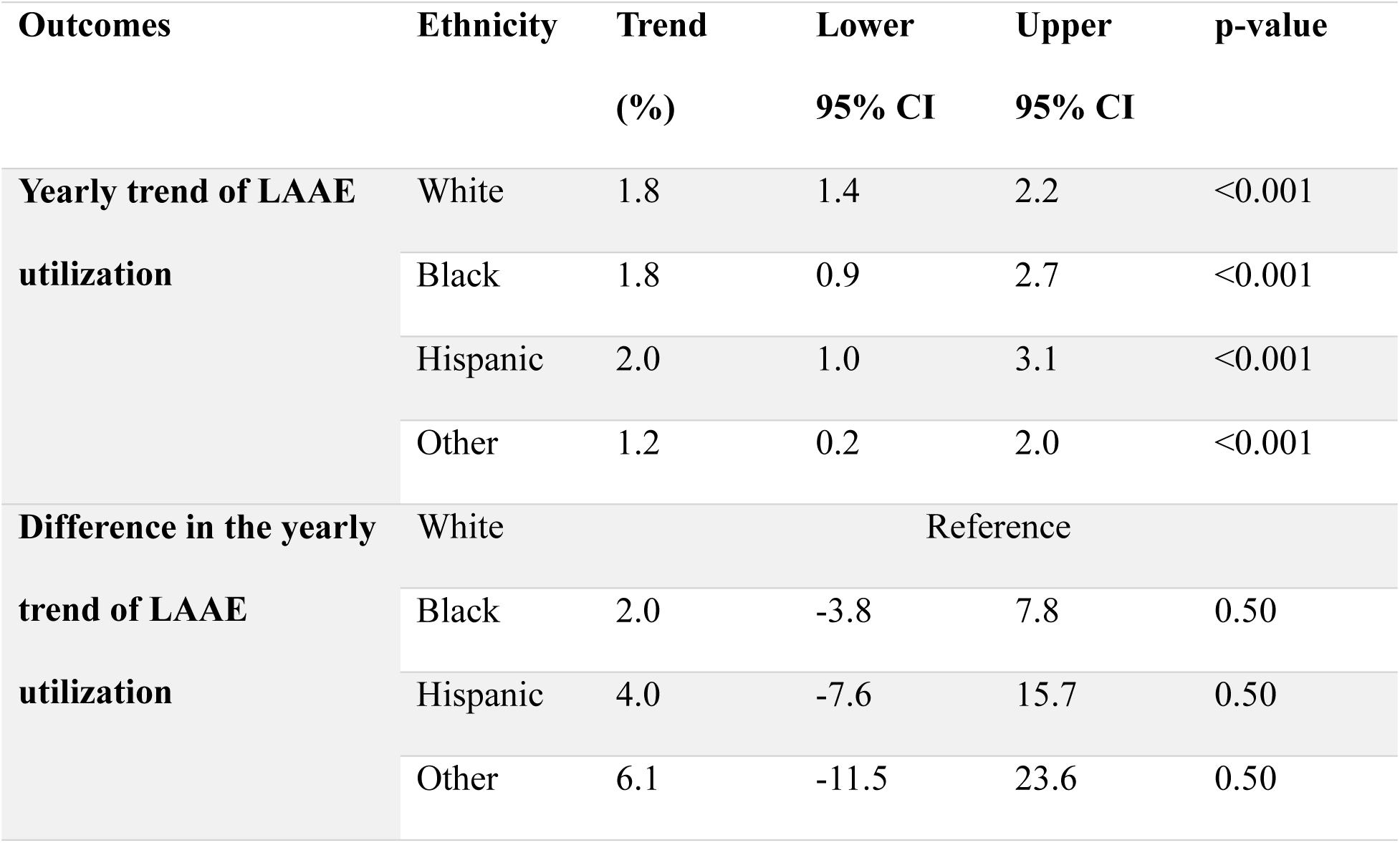
Year on Year trend of LAAE by Race. Difference in yearly trend of concomitant LAAE was calculated using linear regression with an interaction between race and year comparing the yearly utilization between white and non-white races.

## Discussion

This was a retrospective cross-sectional study of over 90,000 patients who underwent concomitant LAAE at the time of cardiac surgery. Our study demonstrated several novel findings: 1) Black and Hispanic patient have lower odds of receiving concomitant LAAE during cardiac surgery compared to White patients; 2) in-hospital mortality and most other clinical outcomes were similar between Black and Hispanic patients compared to White patients; 3) Hospital LOS and total charges were higher in both Black and Hispanic patients compared to white patients; 4) The trend of concomitant LAAE has increased from 2016 through 2020 in all racial/ethnic groups without differences in the trend between White and non-White races.

### LAAE utilization and yearly trend

In our study, non-White patients were less likely to undergo concomitant LAAE during cardiac surgery compared to White patients. These results are similar to Lopez et al. study showing that both Black and Hispanic patients were less likely to receive percutaneous LAA occlusion compared to White patients (aOR 0.45, 95% CI 0.40-0.50 for Black patients and aOR 0.84, 95% CI 0.70-0.99 for Hispanic patients) (18). Similarly, in a study evaluating racial disparities in concomitant LAAE during valve surgery, Black patients were less likely to receive concomitant LAAE during valve surgery (aOR 0.91, 95% CI 0.83-0.99). However, Hispanic patients were equally likely to receive LAAE (aOR 0.99, 95% CI 0.9-1.09) (19). Previous studies have suggested this link being possibly attributed to regional differences in the performance of LAAE, with southern states which underutilize the procedure having a disproportionate number of Black and Hispanic patients (19). However, these disparities persisted in our study even after adjusting for regional differences. Furthermore, these findings are more striking given that most of the clinical outcomes for Black and Hispanic patients were similar compared to White patients. It is to be noted we could not ascertain if there were differences in the proportion of White and non-White patients agreeing to consent for concomitant LAAE during cardiac surgery. Historical distrust and language barriers, as in the case of Black and Hispanic patients could potentially lead to lower proportion of patients consenting to concomitant LAAE (20–23). Overall, there was an increase in the proportion of patients undergoing concomitant LAAE during cardiac surgery in each race/ethnic category. The trend of increase was similar between the White and non-White races suggesting no widening of the gap in the utilization of LAAE during cardiac surgery.

### In-hospital complications

Our study showed no differences in in-hospital mortality and most complications among Black and Hispanic patients compared to White patients. In a study investigating the racial disparities in outcomes for CABG, Black patients had worse postsurgical mortality after CABG compared to white patients and were more likely to undergo CABG at hospitals with the highest mortality compared to white patients (56% vs 47%). However, after adjustment, 30-day mortality was similar between the Black and Hispanic patients (24). In a study based on a veteran’s affairs database, adjusted 30-day mortality after CABG was similar between Black patients and White patients (aOR 1.07, 95% CI 0.84-1.35). However, Black patients did have higher levels of reoperation for bleeding (3.5% vs 2.6 %, p<0.01) but similar rates of stroke or coma (2.9 vs 2.3, p=0.06). Among Hispanic patients, 30-day mortality was lower (aOR 0.70, 95% CI 0.49-0.98), while the rates of stroke/coma and reoperation for bleeding were similar between ( 2.8% vs 2.3%, p=0.24 for stroke/coma and 2.9% vs 2.6%, p=0.43 for reoperation for bleeding) compared to white patients (25). In a study by Alkhouli et al. evaluating the racial disparities in the outcomes of percutaneous LAA occlusion and transcatheter mitral valve repair, there were no differences in the rates of mortality, stroke or pericardial complications between both Black and Hispanic patients compared to White patients (26). In our cohort, both Hispanic and Black patients had similar odds of stroke compared to White patients, but Hispanic patients had higher odds of bleeding complications. It is unclear why Hispanic patients had higher odds of bleeding complications with LAAE but one of the theorized reasons is due the higher burden of liver disease and diabetes in this population, which are known risk factors for bleeding complications.

### LOS of Total hospital charges

Our analysis found that non-White races were more likely to have longer LOS and total hospital charges compared to White patients. The results are most likely due to a higher burden of comorbidities in the non-white races compared to white races. Our results are similar to the results reported by Vincent et al. in an evaluation of racial disparities in percutaneous LAA occlusion where Black patients were more likely to have higher LOS (2.12 ± 3.51 days vs 1.39 ± 1.50 days; P <.001) and higher total hospital charges ($119,317 ± $73,940 vs $113,577 ± $60,984; P = 0.02) compared to White patients (27). Similarly, Khan et al. reported similar results of higher LOS and hospital charges in non-White patients admitted for percutaneous LAA occlusion (28). Overall, these disparities speak to the gap in baseline health status between White and non-White races, which is possibly leading to increased LOS and hospital charges in non-white races with concomitant LAAE.

### Limitations

Our study has several important limitations. 1) As NIS is primarily an administrative database, it can be prone to incorrect coding, thus leading to erroneous conclusions. 2) The study was performed retrospectively and is thus prone to selection bias, as well as the effects of both known and unknown confounders. 3) Given the lack of an ICD-10 code for history of LAAE, we could not confirm that any of our patients did not have their LAA excluded previously. However, we excluded patients who had a history of valvular surgery and CABG. 4) We could not confirm the successful exclusion of the LAA. 5) we could not assess for the use of anticoagulation and thus cannot confirm if there was a difference in the use of anticoagulation between race/ethnicities 6) Our outcomes analysis is limited to inpatient outcomes, and it is possible that our results would be different if there were more longitudinal follow up.

However, despite these limitations, our study provides valuable insight into the disparity in the performance of LAAE based on race and insight on the yearly increase in the trend of LAAE in White, Black, and Hispanic patients.

## Conclusion

Among patients with atrial fibrillation, concomitant LAAE during cardiac surgery is underutilized in Hispanic and Black patients compared to White patients despite mostly similar clinical outcomes. However, there has been an increase in the proportion of patients undergoing concomitant LAAE in all these races from 2016 to 2020.

## Data Availability

All data used for this study is available through HEALTHCARE COST & UTILIZATION PROJECT.

https://hcup-us.ahrq.gov/toolssoftware/comorbidityicd10/comorbidity_icd10.jsp.

## Acknowledgements

None.

## Non-standard Abbreviation and Acronym list

LAAE: Left atrial appendage exclusion.
HCUP: Healthcare Cost and Utilization Project.
NIS: National Inpatient Sample
AHRQ: Agency for Health Care Research and Quality
ICD-10: International Classification of Diseases, tenth revision
CABG: Coronary Artery Bypass Grafting
LAA: Left atrial appendage
AF: Atrial fibrillation

